# Post-acute COVID-19 associated with evidence of bystander T-cell activation and a recurring antibiotic-resistant bacterial pneumonia

**DOI:** 10.1101/2020.09.17.20190033

**Authors:** Michaela Gregorova, Daniel Morse, Tarcisio Brignoli, Joseph Steventon, Fergus Hamilton, Mahableshwar Albur, David Arnold, Matthew Thomas, Alice Halliday, Holly Baum, Christopher Rice, Matthew B. Avison, Andrew D. Davidson, Marianna Santopaolo, Elizabeth Oliver, Anu Goenka, Adam Finn, Linda Wooldridge, Borko Amulic, Rosemary J. Boyton, Daniel M. Altmann, David K. Butler, Claire McMurray, Joanne Stockton, Sam Nicholls, Charles Cooper, Nicholas Loman, Michael J. Cox, Laura Rivino, Ruth C. Massey

## Abstract

Here we describe the case of a COVID-19 patient who developed recurring ventilator-associated pneumonia caused by *Pseudomonas aeruginosa* that acquired increasing levels of antimicrobial resistance (AMR) in response to treatment. Metagenomic analysis revealed the AMR genotype, while immunological analysis revealed massive and escalating levels of T-cell activation. These were both SARS-CoV-2 and *P. aeruginosa* specific, and bystander activated, which may have contributed to this patient’s persistent symptoms and radiological changes.

## Main Text

The COVID-19 pandemic has brought with it the largest ever cohort of patients requiring mechanical ventilation. Complications associated with such severe viral infections are many-fold, and include increased susceptibility to secondary bacterial infections^1,2^, as well as post-acute COVID-19, where patients experience symptoms extending beyond three weeks from the onset of their first COVID-19 symptoms^3^. The first report of secondary infections in COVID-19 patients was from Wuhan in March 2020, where 15% of hospitalized patients developed secondary infections, and of those who did not survive their SARS-CoV-2 infection, 50% had a secondary bacterial infection^1^. Since then many COVID-19 studies reporting secondary infections have been published, with a recent meta-analysis of 24 independent studies that included 3338 patients from five countries reporting that 14.3% of hospitalized COVID-19 patients developed a secondary bacterial infection, which is associated with significant morbidity, mortality and the financial costs associated with prolonged hospitilisation^2^. The incidence of post-acute COVID-19 varies depending on the group of patients considered, with approximately 10% of patients who have tested positive for SARS-CoV-2 virus remaining unwell beyond three weeks^3^. However, this can be as high as 74% when hospitalised patients are considered, where symptoms include breathlessness and excessive fatigue, with abnormal radiological features reported in 12% of this cohort^4^.

The DISCOVER study (DIagnostic and Severity markers of COVID-19 to Enable Rapid triage, REC: 20/YH/0121) was established in March 2020 to collect and analyze longitudinal samples from COVID-19 patients. One study participant, an otherwise healthy male between 45 and 55 years of age presented to hospital with Type-1 respiratory failure, 20 days after he tested positive for SARS-CoV-2 by RT-PCR. At the time of testing he was asymptomatic (tested as a house-hold contact of a health-care worker), and this represents Day 1 on the time-line presented in fig. 1. He became symptomatic for COVID-19 13 days after this and his health declined over the following week. Upon admission to hospital a chest X-ray was taken, and he was admitted to the ICU where he was mechanically ventilated (fig. 1). An RT-PCR test on an endotracheal sample collected at this time did not detect SARS-CoV-2 suggesting that he had cleared the viral infection.

**Fig. 1:**
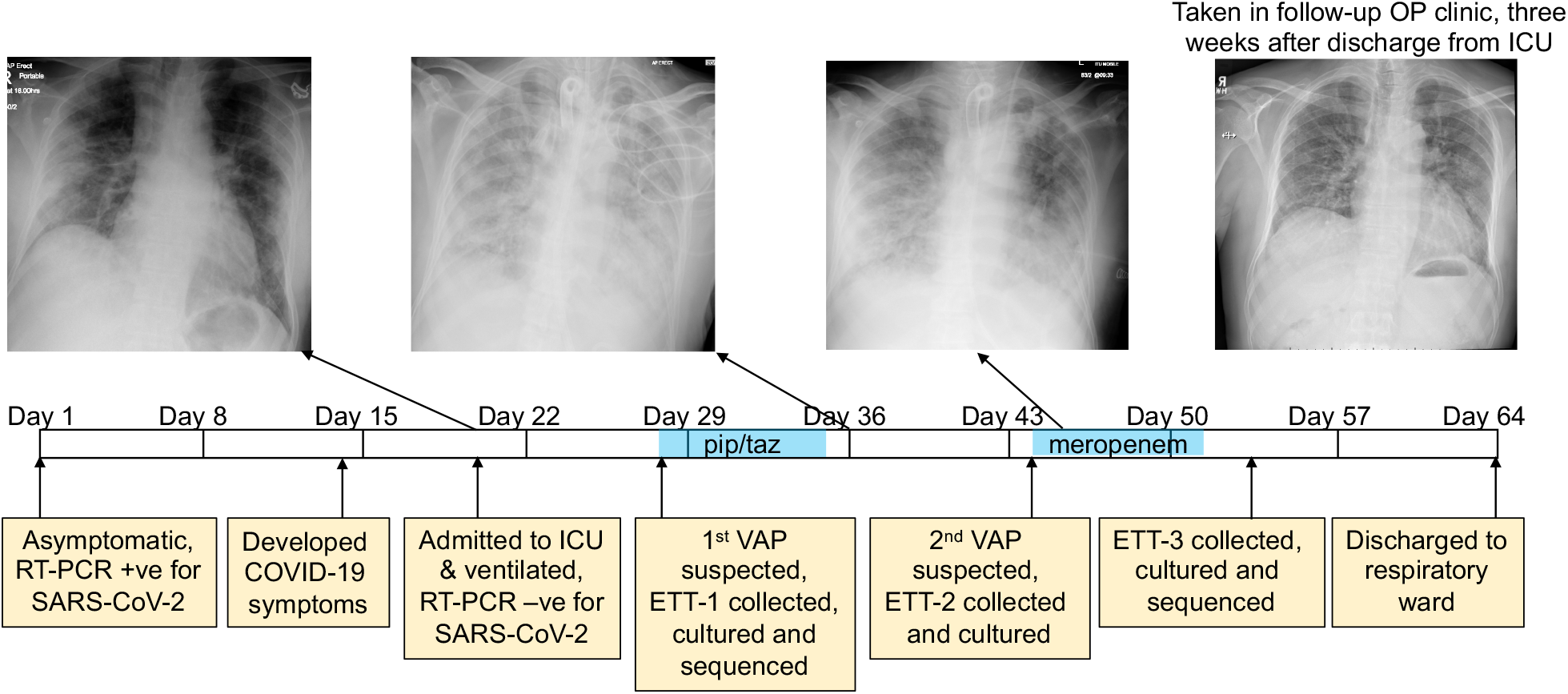
The development of a recurring ventilator-associated pneumonia (VAP) by a COVID-19 patient. A clinical time-line is presented from the point at which the patient tested positive for SARSCoV-2 (day 1), through to his discharge from the ICU (day 64). Noteworthy clinical features are indicated in the yellow boxes below the time-line. The three x-rays taken during the patient’s time in the ICU are presented, as is a later follow-up x-ray taken in an out-patient (OP) clinic three weeks after discharge. The time during which the antibiotics piperacillin and tazobactam (pip/taz) and meropenem were administered to the patient to treat the VAP are indicated in blue. The points at which endotracheal tube aspirates (ETT) were collected and the subsequent analysis of these also are indicated and described.

After a week in the ICU he was diagnosed with ventilator-associated pneumonia (VAP) based on clinical, radiological, biochemical and microbiological parameters. An antibiotic sensitive *Pseudomonas aeruginosa* strain was cultured from an aspirate collected from his endotracheal tube (ETT-1, fig. 1). He was prescribed a seven-day course of piperacillin-tazobactam (pip/taz) (4.5g every six hours), and clinically recovered from this bacterial infection. Eight days after finishing this first course of antibiotics his VAP recurred, and a pip/taz resistant *P. aeruginosa* was cultured from ETT-2 (minimum inhibitory concentration (MIC) >16mg/l). He was prescribed a seven-day course of meropenem (1g every 8 hours) and showed signs of clinical improvement. There were no further concerns about a bacterial infection during his ICU stay although a third *P. aeruginosa* was cultured from a sample (ETT-3) collected two days after he completed his course of meropenem, that was resistant to both pip/taz and meropenem (MICs >16 mg/l and >8mg/l respectively). His health continued to improve, and he was discharged to the respiratory ward a week later. The patient attended a follow-up clinic 2-weeks after hospital discharge where he reported on-going symptoms such as breathlessness and myalgia. A chest X-ray demonstrated persistent ground-glass opacities throughout the lungs (fig. 1) and there was a significant drop in his oxygen saturations after mild exertion to 84%.

To understand the cellular and molecular dynamics of this post-acute COVID-19 case from the perspective of both the pathogen and patient’s immune response, longitudinal respiratory and blood samples were collected and analyzed with a view to identifying early diagnostic biomarkers of infection onset and potential opportunities for immunotherapeutic intervention.

Metagenomics was used to characterise in depth the composition and genomic features of the bacteria present in this patient’s lower respiratory tract from when he first developed VAP (ETT-1, fig. 1) to when he had recovered from his second VAP (ETT-3, fig. 1). We extracted the entire genetic material from 500µl of ETTs 1 and 3 with no bacterial enrichment or human DNA depletion steps, and to ensure data for the bacterial component of the samples was generated, these were sequenced on a PromethION (Oxford Nanopore technology). From ETT-1 whole genomes for both *P. aeruginosa* and *Staphylococcus haemolyticus* (fig. 2) were generated. The *P. aeruginosa* isolate corresponded to multi-locus sequence type ST253^5^, a world-wide clone frequently associated with AMR epidemics^6^. While the *S. haemolyticus* isolate corresponded to ST10 (albeit with a new allele for the SH1431 locus^5^) and carried the methicillin resistance conferring SCC*mec* element^7^. Coagulase negative staphylococci such as *S. haemolyticus* are not recognised as lung pathogens and therefore typically ignored in a diagnostic laboratory as probable contaminants from ETT samples. That we generated a whole genome for this from this sample with no enrichment suggests it was present in a significant abundance, although the clinical relevance of this remains to be determined.

**Fig. 2:**
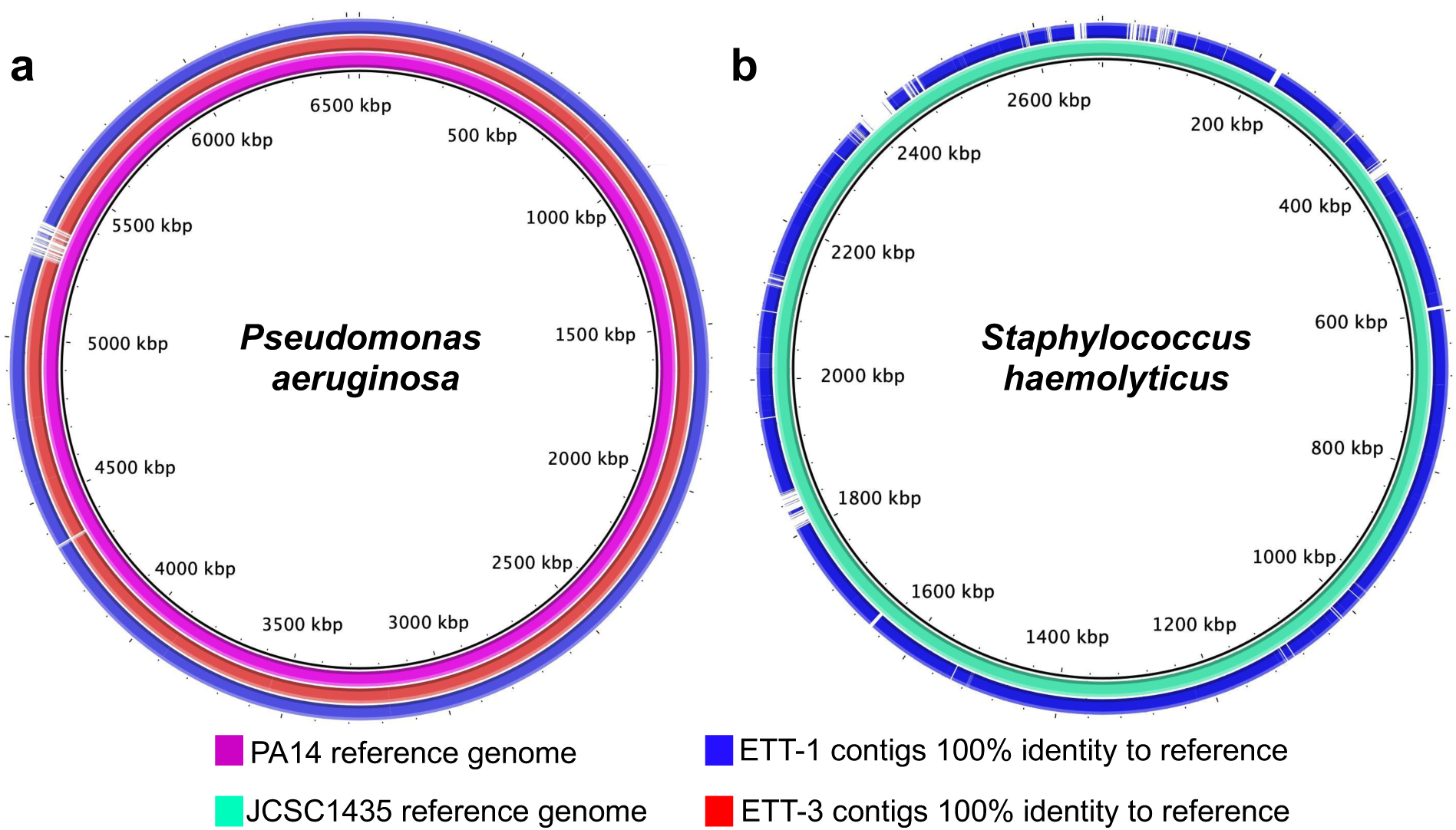
Direct metagenomic analysis of respiratory samples from a COVID-19 patient who developed a recurring VAP. (**a**) The sequence data were aligned to the genome of a *P. aeruginosa* reference strain PA14, represented in pink. The blue genome represents the regions within the *P. aeruginosa* genome in ETT-1 that has 100% identify to the PA14 reference genome. The red genome represents the regions within the *P. aeruginosa* genome in ETT-3 that has 100% identify to the PA14 reference genome. (**b**) The sequence data were aligned to the genome of a *S. haemolyticus* reference strain JCSC1435, represented in green. The blue genome represents the regions within the *S. haemolyticus* genome in ETT-1 that has 100% identify to the JCSC1435 reference genome. Gaps in the genomes indicate regions present in the reference that are absent from that in the ETTs.

Resistance to beta-lactam antibiotics by *P. aeruginosa* can be multifactorial and includes the acquisition of single nucleotide polymorphisms (SNPs) in efflux and porin genes that affect the passage of the antibiotic into and out of the bacterial cell^8^. Analysis of the sequence data from ETT-3 generated a whole genome for *P. aeruginosa*, again corresponding to ST253, that aligned with >99% identify to the genome from ETT-1. There were however two noteworthy SNPs in this later sample that explain the increased AMR of this isolate. The *mexR* gene encodes a repressor of the MexAB antibiotic efflux pump, and we found a SNP in *mexR* that converts a key amino-acid (Arg91-Gln) in the DNA binding domain of the protein, that would likely result in the de-repression of this efflux system^9^. A second SNP introduced a premature stop codon in the *oprD* porin encoding gene (Tyr120-stop), that encodes a protein with a well-established role in the entry of meropenem into the bacterial cell^10^.

Having recovered from both a viral and a recurring bacterial infection, we also sought to analyse the kinetics of the patient’s innate and adaptive immune response across his time in the ICU (at days 23, 28, 38, 45 and 58 from testing positive for SARS-CoV2, fig. 1). Robust activation of a broad range of immune cell subsets was revealed by flow cytometry, when compared to healthy controls. What was particularly striking was the level of activation and proliferation of CD4+, CD8+ and TCR-gd T-cells, that appeared to wane between days 23-28 but were subsequently boosted after day 28, concomitant with the onset of the secondary bacterial infection (Figures 3 a-c, Supplementary Figure S1). From day 28 a robust and steady increase in the activation and proliferation of conventional and TCR-gd-T-cells was evident with approximately 20% and 40% of total CD4+ and CD8+ T-cells co-expressing the activation markers HLA-DR/CD38, respectively. A similar steady increase in activation levels, albeit at lower magnitudes, was observed for Natural Killer (NK) CD56^dim^ and CD56^bright^ cells (Figure 3d, Supplementary Figure S1). Similar perturbations in the frequency and activation phenotypes of monocytes, blood monocyte-derived macrophages as well as neutrophils could be detected from day 28 onwards (Supplementary Figure S2). A robust IgG response to both SARS-CoV-2 and *P. aeruginosa* antigen was also detected in this patient (Supplementary Fig S4).

Given the scale of the cellular response of this patient, we investigated whether the large expansions of CD4+ and CD8+ T-cells were due to a T-cell response targeting SARS-CoV-2, the secondary bacterial infection or to bystander T-cell activation^11,12^. We included the analysis of bystander activation, which is T-cell receptor-independent and cytokine-mediated, as it is known to occur during other types of acute viral infection^11,12^. The role of this type of T-cells activation in the recovery of patients remains largely unclear, as these cells can participate in protective immunity towards the virus but can also contribute to tissue damage^11–14^. We performed a brief stimulation of the patient’s PBMCs, with or without overlapping peptides spanning the sequences of SARS-CoV-2 proteins (spike, membrane (M) and nucleoprotein (N)); of an immunodominant *P. aeruginosa* antigen (OprF); as well as an immunodominant human Cytomegalovirus protein (HCMV pp65) as an indication of non-T Cell Receptor (TCR) driven bystander activation. This was followed by intracellular cytokine staining to detect production of IFN-g and TNF-a by the activated cells. Following the encounter with specific peptides we observed a robust CD4+ T-cell response to SARS-CoV-2 which decreased from day 23 to day 58, and a more modest SARS-CoV-2-specific CD8+ T-cell response. This is in line with published work, suggesting higher magnitudes of SARS-CoV-2-specific CD4+ versus CD8+ T-cells in severe COVID-19 patients^15^, as well as consistent detection of virus-specific CD4+ T cells in recovered patients^16^ (Figure 1e-f, Supplementary Figure S3). CD4+ and CD8+ T-cell responses targeting *P. aeruginosa* peptides could also be detected and appeared to increase over time concomitant with the onset of the recurring bacterial infection (Figures 1e-f). However, that the number of T-cells responding specifically to SARS-CoV-2 and *P. aeruginosa* represent only a fraction of those activated suggests that many of those detected in the patient’s blood may be bystander activated, which is supported by the increased magnitude of CMV-specific T-cell responses we detected over time (Figures 1e-f). Whether the immune perturbations associated here with a case of post-acute COVID-19 are attributable to the secondary bacterial infection is currently under investigation.

In this case report we demonstrate the potential of the application of multidisciplinary technologies to longitudinally-collected patient samples to define the complex dynamics of patient-pathogen-therapy interactions in real-time. Metagenomics directly applied to respiratory samples facilitated the identification of the SNPs responsible for the AMR phenotype of the later *P. aeruginosa* isolate. However, the most striking feature of this COVID-19 case was the escalating number of circulating activated T-cells more than two months after testing positive for SARS-CoV-2, and six weeks after clearing the viral infection. While we could attribute some of this to the recurring bacterial infection, the scale of the activation considered alongside our evidence of increased frequencies of T-cells specific for unrelated antigens, suggests there may be a significant amount of bystander activation. This could play a critical role in the severity of illness and the longer-term complications associated with the development of post-acute COVID-19 experienced by this patient^17^. Given the recent appreciation for the role of corticosteroids in reducing the risk of death following SARS-CoV-2 infection by 20%^16^, this case suggests that targeting its use to patients with immunological responses as described here, may also improve their long-term recovery.

**Fig. 3:**
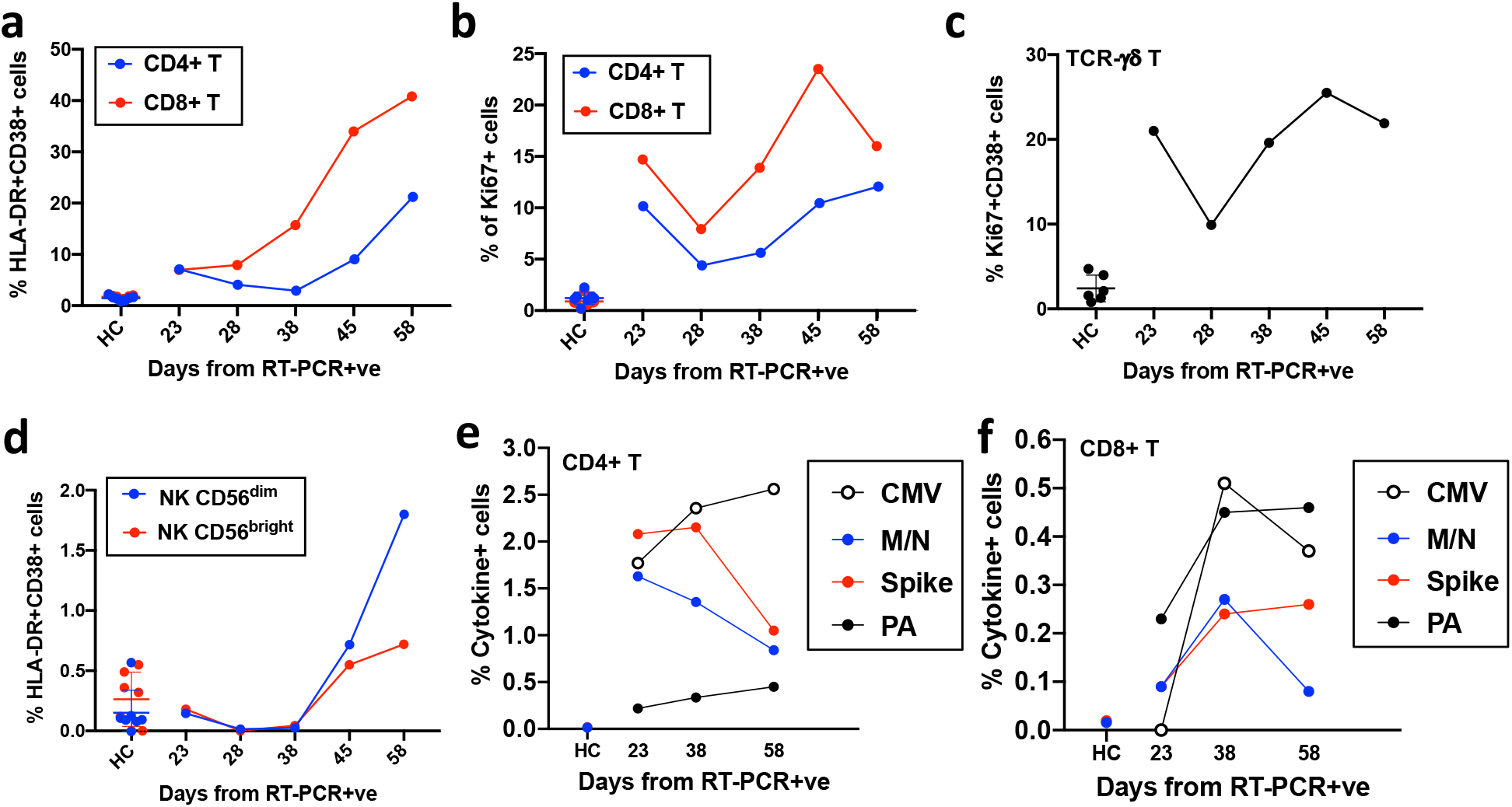
Kinetics of the adaptive and innate immune response in a COVID-19 patient during ICU treatment. **(a-b)** T-cell activation and proliferation measured as co-expression of the activation markers HLA-DR/CD38 and Ki67, respectively, is shown for CD4+ and CD8+ T-cells from the COVID-19 patient and healthy controls (HC, n = 7). **(c)** Activation and proliferation of TCR-gd T cells measured as co-expression of Ki67 and CD38 is shown for the COVID-19 patient and for healthy controls. **(d)** Activation of NK CD56^dim^ and CD56^bright^ cells measured as coexpression of the activation markers HLA-DR and CD38 is shown for the COVID-19 patient and for healthy controls. **(e-f)** T cell specificity is assessed in CD4+ (e) and CD8+ (f) T-cells by intracellular cytokine staining after a brief stimulation of PBMCs with HCMV pp65 (CMV), SARSCoV-2 membrane, nucleoprotein, spike (M/N and Spike, respectively) and *Pseudomonas aeruginosa* OprF (PA) peptides. Responses are shown as % of IFN-g and/or TNF-a+ cells in CD4+ or CD8+ T cells after subtraction of the negative control in the patient and in a age-matched healthy control (spike and M/N). Gating strategies and dot plots are included in Supp. Figures S1 and S3.

## Methods

### Patient Recruitment

The patient was enrolled onto the DISCOVER study (Diagnostic and Severity markers of COVID-19 to Enable Rapid triage study), a single centre prospective study recruiting consecutive patients admitted with COVID-19, from 30.03.2020 until present (Ethics approval via South Yorkshire REC: 20/YH/0121, CRN approval no: 45469). Blood/serum samples from pre-pandemic healthy controls and asymptomatic healthy controls were obtained under the Bristol Biobank (NHS Research Ethics Committee approval ref 14/WA/1253).

### Clinical Microbiology

All ETT samples were processed in the Severn Infection Services laboratory as per standard operating procedures. Antibiotic susceptibility testing was performed by either disk diffusion (according to EUCAST version 9 guidance^18^) or on a VITEK 2 machine (BioMeriux, France).

### SARS-CoV-2 test

SARS-CoV-2 test was performed by an in-house RT-PCR at the regional South West Public Health England Regional Virology laboratory, utilising a PHE approved assay at the time of testing.

### DNA extractions

DNA from the endotrachaeal aspirates were extracted using the CTAB/Phenol:Chloroform:Isoamyl alcohol and bead-beating approach of Griffiths et al^19^ with modifications of DeAngelis et al^20^. Phase lock gel tubes (ThermoFisher) and linear polyacrylamide (Sigma) were included to increase nucleic acid yields and total DNA was resuspended in 50 µl of DNase/RNase free water before storage at −20°C.

### DNA sequencing

Samples were prepared for sequencing using SQK-LSK109 kit (Oxford Nanopore) with 1µg DNA starting input as per manufacture’s protocol. Briefly 1µg of DNA was end repaired and a tailed using NEBNext Ultra II module E7546 (3.5µl End Repair Buffer, 2ul FFPE repair mix, 3.5 µl Ultra II end-prep reaction buffer and 3µl of Ultra II end-prep enzyme mix to 1µg DNA in a total of reaction volume of 60µl). This was incubated at 20°C for 5 min followed by 65°C for 5 min. Clean-up was performed using AMPure XP beads (Beckman Coulter) in a 1X ratio.

Adaptors were ligated by adding 5µl adaptor mix (Oxford Nanopore) 25µl ligation buffer (Oxford Nanopore) and 10µl Quick T4 ligase (NEB Module MO202). Following a 20-minute incubation at room temperature the adaptor ligated DNA was cleaned using AMPure beads in a 0.8 X ratio and washed using Long Fragment Buffer (Oxford Nanopore) before eluting in 25µl of elution buffer (Oxford Nanopore). Final quantification by fluorometry (Qubit) was performed and 300ng DNA prepared for sequencing according to the manufacturer’s instructions (Oxford Nanopore). Sequencing was performed on a PromethION R9.4.1 flow cell (FLO-PRO002) and run for 48 hours using live basecalling, files were outputted in Fast5 and Fastq format. Human-filtered sequencing data for this study have been deposited in the European Nucleotide Archive (ENA) at EMBL-EBI under accession PRJEB40239.

### Bioinformatics

Bioinformatics was orchestrated using reticulatus (https://github.com/SamStudio8/reticulatus/) with configuration: Flye (v2.6) metagenomic assembly^21^, Racon polishing (v1.4.9 + GPU, 2 rounds)^22^ and Medaka polishing (v0.8.0 + GPU, 1 round). Contigs were assigned to a taxon by Kraken 2^23^. SNP differences between the *P. aeruginosa* contigs for ETT-1 and ETT-3 was determined using *Mauve*^24^. Contigs were loaded into BRIG (v0.95)^24^ as concentric rings and compared against the PA14 or JCSC1435 reference genomes using blastn (ncbi-blast 2.10.1+)^25^. MLST types were identified by mapping contigs against *P. aeruginosa* or *S. haemolyticus* according to the schemes held in the pubmlst databases^4^.

### PBMC isolation

Blood samples were collected from the COVID-19 patient in EDTA vacutainer tubes and PBMCs isolated from peripheral blood by Ficoll gradient purification and cryopreserved. Healthy donor PBMCs were obtained from Bristol Biobank (REC: 14/WA/1253).

### Synthetic peptides

15-mer peptides overlapping by 10 amino acids and spanning the sequences of SARS-CoV-2 spike (Accession Number: NC_045512.2, Protein ID: YP_009724390.1) and HCMV pp65 (AD169 strain) were purchased from Mimotopes (Australia). The purity of the peptides was >80% (Spike) or >70% (pp65) and peptides were dissolved as described previously^10^. SARS-CoV-2 M and N Peptivator peptide pools were purchased from Miltenyi. An OprF (PA1777) peptide library comprising 20-mer peptides overlapping by 10 amino acids was synthesized by GL Biochem Ltd., Shanghai, China^26^.

### Flow cytometry staining and PBMC stimulation

PBMCs were thawed and either stained ex vivo or stimulated in AIMV 2% FCS with or without peptide pools from SARS-CoV2 spike, M, N, HCMV pp65 (all 1µg/ml), OrpF PA (10µg/ml) or with PMA/iono (PMA 10 ng/ml, Iono 100 ng/ml, Sigma Aldrich) for 5 hours at 37 ºC in the presence of brefeldin A (BD, 5µg/ml). To assess degranulation, CD107a FITC antibody was added to the cells at the beginning of the stimulation. Cells were stained with a viability dye Zombie Aqua (Biolegend) for 10 min at room temperature and with antibodies targeting surface markers (20 min 4ºC, diluted in PBS (HyClone) 1% BSA (Sigma Aldrich)). Cells were fixed for 45 min/overnight in eBioscience Foxp3/Transcription factor fixation/permeabilization buffer (Invitrogen) and intracellular staining was performed using eBioscience Foxp3/Transcription factor permeabilization buffer (Invitrogen) for Ki67 or intracellular cytokines (30 min on ice). Cells were acquired on a BD Fortessa X20 and data analyzed using FlowJo software v10.7. A complete list of antibodies is included in (Supplementary Table S1.

## Data Availability

All data is freely available

## Acknowledgements

We would like to thank Kapil Gupta and Imre Berger for kindly providing us the Spike protein, Natalie Di Bartolo and Ashley Toye for kindly providing us the N protein, both used for the SARS-CoV-2 serology work. We would also like to thank Keith Jolley and the Bristol University UNCOVER team for helpful discussions during the execution of this work and preparation of the manuscript. This work was supported by donations to Southmead Hospital Charity (Registered Charity Number: 1055900), by the Wellcome Trust (reference number: 212258/Z/18/Z) and by the Elizabeth Blackwell Institute, University of Bristol, with funding from the University’s alumni and friends. D.K.B. is supported by a Cystic Fibrosis Trust PhD studentship (CF Trust SRC 015). R.J.B. and D.M.A. are supported by UKRI (MR/S019553/1 & MR/R02622X/1)

